# Artificial Intelligence in Medical Imaging With Emphasis on Generative and Foundation-Based Methods: A Bibliometric Analysis of Global and United Kingdom Research, 2017-2025

**DOI:** 10.64898/2026.06.26.26356684

**Authors:** Jatin Sridhar Naidu, Vasanth Baskaradoss

## Abstract

**Background:** Artificial intelligence (AI), including generative and foundation-based methods, has rapidly expanded within medical imaging research. However, the structure, citation impact, collaboration patterns, and thematic orientation of national research ecosystems remain incompletely characterised.

**Objectives:** To evaluate global research trends in AI applied to medical imaging between 2017 and 2025, with detailed analysis of United Kingdom (UK)-affiliated output, citation performance, collaboration structure, funding landscape, and thematic evolution, with emphasis on generative and foundation-based methodologies.

**Materials and Methods:** A bibliometric analysis of Scopus-indexed publications (2017-2025) was performed using a predefined search strategy targeting AI and medical imaging concepts, with emphasis on generative and foundation-based terms. Records were analysed globally and filtered for UK affiliation. Descriptive indicators including total publications (TP), total citations (TC), citations per paper (CPP), and year-on-year growth were calculated. Co-authorship and keyword co-occurrence networks were generated using VOSviewer (v1.6.19).

**Results:** A total of 13,452 publications were identified globally (194,650 citations; global CPP 14.47), of which 889 (6.61%) were UK-affiliated. The UK ranked fourth by publication volume yet demonstrated higher citation efficiency (CPP 21.00) than several higher-volume countries. UK output increased approximately 18-fold between 2017 and 2025, with evidence of a citation-lag effect in recent years. Research activity was concentrated within a small number of institutions accounting for nearly half of national output, although citation impact varied independently of volume. Journal-dominant dissemination was associated with higher average citation impact compared with conference-centric models. Keyword analysis identified three principal thematic clusters: generative/deep learning methodologies, MRI- and diffusion-focused applications, and broader diagnostic imaging workflows. Highly cited publications were initially dominated by generative adversarial network–based reconstruction and synthesis, with recent rapid citation growth observed in diffusion and foundation-model architectures.

**Conclusion:** UK-affiliated research represents a rapidly expanding and highly cited component of the global AI medical imaging literature, with increasing emphasis on generative, diffusion-based, and foundation-model approaches. These findings provide a reproducible bibliometric baseline for monitoring research activity, collaboration patterns, and potential translational priorities, while recognising that citation-based indicators do not directly measure clinical implementation, methodological quality, or real-world impact.

## INTRODUCTION

Artificial intelligence (AI) encompasses computational methods that enable machines to perform tasks traditionally requiring human intelligence, including pattern recognition, decision-making, and data interpretation.^[1]^ Advances in machine learning and deep learning have substantially expanded the capacity of AI systems to learn complex representations from large datasets, driving rapid research activity across many areas of healthcare. Within medical imaging, AI research has explored applications in image acquisition, reconstruction, segmentation, synthesis, triage, and interpretation, with the potential to support diagnostic efficiency and consistency.^[2]^ These developments have positioned AI as a central area of investigation in radiology and other imaging-intensive specialties.

In recent years, generative artificial intelligence has emerged as a particularly influential class of methods within medical imaging. Techniques such as generative adversarial networks (GANs), diffusion models, variational autoencoders, and foundation-based architectures enable the synthesis and transformation of images, offering solutions to longstanding challenges including limited annotated data, image noise, motion artefacts, and cross-modality translation.^[3]^ Generative models have been applied to diverse tasks such as accelerated magnetic resonance imaging reconstruction, image enhancement, data augmentation, anomaly detection, and multimodal image synthesis.^[4,5]^ Collectively, these approaches extend beyond traditional discriminative modelling and support more comprehensive manipulation and understanding of imaging data.

The rapid growth of generative and foundation-based AI research has produced a large, heterogeneous body of literature spanning clinical journals, engineering venues, and conference proceedings.^[6]^ As this field expands, it becomes increasingly important to understand how research activity is distributed, which countries and institutions contribute most substantially, how collaborative networks are structured, and which thematic areas dominate scholarly output. Such insights are essential for informing research strategy, funding allocation, and translational planning.^[7]^

Bibliometric analysis provides a systematic and reproducible framework for quantitatively assessing research output, citation impact, collaboration patterns, and thematic evolution within large scientific corpora. By applying mathematical and statistical techniques to bibliographic metadata, bibliometric methods enable the identification of influential publications, emerging research topics, and structural relationships among authors, institutions, and countries.^[8]^ Previous bibliometric studies have examined artificial intelligence in medical imaging at a global level or within specific modalities and clinical applications.^[9–11]^ However, most have focused on broad AI techniques without detailed attention to generative methodologies or to national research ecosystems.

The United Kingdom represents a particularly relevant context for such an analysis. The UK hosts a strong academic medical imaging community, maintains a nationally integrated healthcare system, and has articulated strategic priorities for responsible AI development and deployment in healthcare.^[12,13]^ These features position the UK as both a major contributor to methodological innovation and a relevant context for translational research and evaluation. Nevertheless, a comprehensive bibliometric characterisation of UK-affiliated research in AI applied to medical imaging, with emphasis on generative and foundation-based methods, has not been previously reported.

Accordingly, the present study aims to conduct a bibliometric analysis of United Kingdom-affiliated research in artificial intelligence applied to medical imaging between 2017 and 2025, with particular emphasis on generative and foundation-based methods. By examining publication trends, citation impact, country and institutional contributions, collaboration networks, and keyword co-occurrence patterns, this study seeks to map the structure and evolution of this research domain and to contextualise the UK contribution within the global literature. These findings provide an empirical baseline to support future research planning and policy discussion, while recognising that bibliometric indicators do not directly establish clinical translation or implementation.

## METHODS

### Data Source and Search Strategy

The Scopus database was used to identify relevant publications. Scopus was selected because of its broad coverage of biomedical, engineering, computer science, and conference proceedings literature, which are central dissemination routes for medical imaging AI research. Searches were performed in the TITLE-ABS-KEY fields. A comprehensive search strategy targeting artificial intelligence and medical imaging concepts was applied for the period 2017-2025, with results filtered to records listing at least one United Kingdom-affiliated author. Although the search strategy prioritised generative and foundation-based artificial intelligence techniques (e.g., generative adversarial networks, diffusion models, and large language models), broader artificial intelligence terms were intentionally included to allow contextualisation of generative methods within the wider medical imaging AI literature. The complete search string is provided below:

TITLE-ABS-KEY(“generative AI” OR “generative artificial intelligence” OR “generative adversarial network*” OR GAN OR “diffusion model*” OR “foundation model*” OR “large language model*” OR ChatGPT OR “Gemini AI”) AND TITLE-ABS-KEY(“medical imag*” OR “diagnostic imaging” OR radiolog* OR CT OR MRI OR ultrasound OR “computed tomography” OR “magnetic resonance”) AND (LIMIT-TO (AFFILCOUNTRY, “United Kingdom”)))

The same search, without country restriction, was used to derive global comparator metrics. The search was executed and the dataset exported on 6 February 2026. As Scopus is continuously updated, rerunning the identical query at a later date may yield slightly different record counts. All analyses were performed on the dataset exported on 6 February 2026.

### Data Extraction and Analysis

Bibliographic information, including publication year, document type, author affiliations, citation counts, funding details, and keywords, was exported from Scopus. Descriptive bibliometric indicators were calculated, including total publications, total citations, citations per paper (CPP), and year-on-year (YoY) growth of UK publications, calculated as [(UK TPIZ − UK TPIZ₋₁) ÷ UK TPIZ₋₁] × 100. United Kingdom affiliation was defined as the presence of at least one UK-based author on the publication; first or corresponding author status was not required.

Keyword co-occurrence and co-authorship analyses were performed using VOSviewer (v1.6.19). Full counting was applied. Generic indexing terms (e.g., “article,” “human,” “humans”) were excluded prior to keyword analysis. For author co-authorship mapping, a maximum of 25 authors per document and a minimum of three documents per author were required for inclusion; from the eligible set, the top 50 authors by total link strength were selected for network construction. For keyword analyses, a minimum occurrence threshold of 10 was applied, and the top 40 keywords meeting this threshold were retained for network construction. Network visualisation thresholds were applied to enhance interpretability, and clusters were identified algorithmically based on co-occurrence and collaboration strength.

### Inclusion and exclusion criteria

Publications were eligible for inclusion if they were indexed in the Scopus database between 2017 and 2025, contained terms related to artificial intelligence and medical imaging within the title, abstract, or keywords, and listed at least one United Kingdom-affiliated author. All document types indexed in Scopus were considered eligible.

Following database retrieval, records were verified for relevance to medical imaging and artificial intelligence–based methodologies and for completeness of bibliographic metadata. Records that were unrelated to the defined scope or contained insufficient metadata were excluded.

## RESULTS

### 1. Overall picture

The research output from 2017 to 2025 showed a marked upward trajectory in global and United Kingdom (UK)-affiliated publishing activity in artificial intelligence (AI) applied to medical imaging. The global output comprised 13,452 publications, accruing 194,650 citations and corresponding to a global citations-per-paper (CPP) value of 14.47. Annual global output increased from 131 publications in 2017 to 4,910 publications in 2025, demonstrating sustained expansion across the study period.

Country-level publication and citation metrics are presented in Table 1 and Figure 1. China contributed the largest number of publications (4,008; 29.79%), followed by the United States (3,249; 24.15%), India (1,480; 11.00%), and the United Kingdom (889; 6.61%). The United States accrued the highest total citation count (73,310; CPP 22.56), followed by China (52,130; CPP 13.00) and the United Kingdom (18,672; CPP 21.00). Among the top 10 countries by publication volume, Canada had the highest CPP (28.07), followed by Australia (25.25), the United States (22.56), and the United Kingdom (21.00).

**Figure 1.**
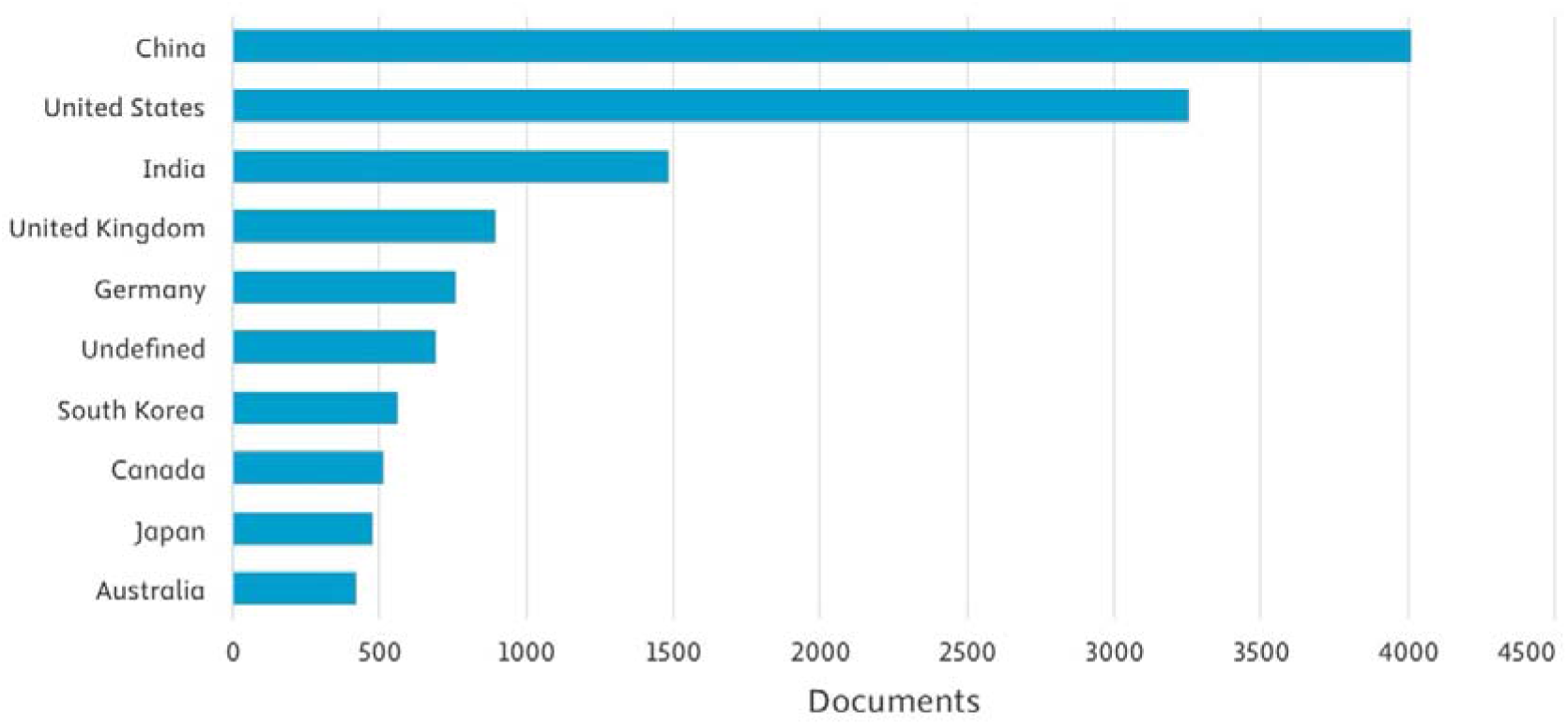
Country-wise distribution of global research output in artificial intelligence applied to medical imaging. Bars represent total publications per country; “Undefined” denotes records with missing or non-specific country affiliation.

**Table 1.** Contribution of the top 10 countries to global research output in medical imaging, with comparison of publication volume and citation impact metrics relative to the United Kingdom. Metrics include total publications (TP), total citations (TC), citations per paper (CPP), and percentage contribution to total output (TP%). *Undefined denotes records with missing or non-specific country affiliation.

| S. No | Name of Country | TP | TC | CPP | TP% |
| --- | --- | --- | --- | --- | --- |
| 1 | China | 4008 | 52130 | 13.00 | 29.79 |
| 2 | United States | 3249 | 73310 | 22.56 | 24.15 |
| 3 | India | 1480 | 11155 | 7.54 | 11.00 |
| 4 | United Kingdom | 889 | 18672 | 21.00 | 6.61 |
| 5 | Germany | 758 | 11208 | 14.78 | 5.64 |
| 6 | Undefined* | 689 | 845 | 1.23 | 5.12 |
| 7 | South Korea | 559 | 10612 | 18.98 | 4.15 |
| 8 | Canada | 513 | 14404 | 28.07 | 3.81 |
| 9 | Japan | 472 | 5999 | 12.71 | 3.51 |
| 10 | Australia | 419 | 10576 | 25.25 | 3.11 |
| TP: Total papers, TC: Total citations, CPP: Citations per paper |  |  |  |  |  |

**Figure 2.**
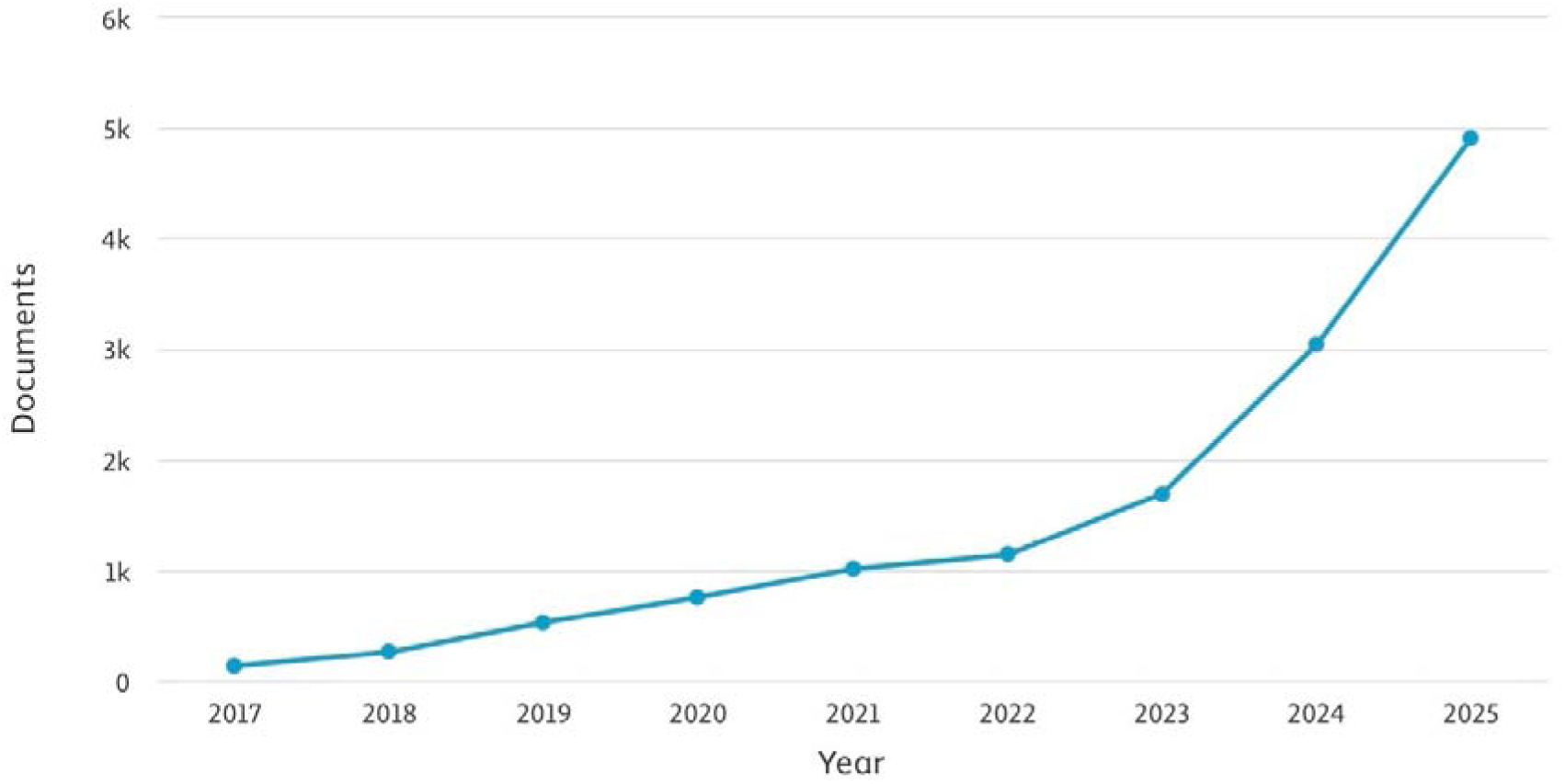
Annual global publication trends in artificial intelligence applied to medical imaging (2017-2025). The line plot shows the number of Scopus-indexed publications per year.

This pattern shows that publication volume and citation efficiency were not directly aligned. Although China produced the largest number of papers, several lower-volume countries, including Canada, Australia, and the United Kingdom, had higher CPP values. The United Kingdom therefore occupied a relatively strong position in citation visibility, ranking fourth by publication volume but achieving higher CPP than several higher-output countries.

### 2. United Kingdom research output and citation profile

Annual trends in global and UK-affiliated output are shown in Table 2. UK-affiliated output increased from 17 publications in 2017 to 305 publications in 2025, representing an approximately 18-fold increase. This growth occurred alongside rapid global expansion, with global output rising from 131 to 4,910 publications over the same period. The UK share of global output was highest in 2017 (12.98%) and subsequently remained at approximately 6-7% for most years from 2019 onwards. In absolute terms, however, UK-affiliated activity increased substantially, with 34.32% of all UK-affiliated publications in the dataset published in 2025.

**Table 2.** Annual trends in global and United Kingdom-affiliated research output in artificial intelligence in medical imaging (2017-2025). The table summarises total publications (TP) globally and from the United Kingdom, the UK share of global output (TP of Global, %), and the temporal distribution of UK publications across the study period (TP of UK, %). Citation impact is reported as total citations (TC) and citations per paper, with year-on-year (YoY) growth reflecting annual changes in UK publication volume.

| Period | Global | UK |  |  |  |  |  |
| --- | --- | --- | --- | --- | --- | --- | --- |
|  | TP | TP | TP of Global (%) | TP of UK (%) | TC | CPP | YoY Growth (%) |
| 2017 | 131 | 17 | 12.98 | 1.91 | 1098 | 64.59 | - |
| 2018 | 256 | 20 | 7.81 | 2.25 | 1947 | 97.35 | 17.65 |
| 2019 | 525 | 35 | 6.67 | 3.94 | 1503 | 42.94 | 75.00 |
| 2020 | 753 | 48 | 6.37 | 5.40 | 2472 | 51.50 | 37.14 |
| 2021 | 1011 | 69 | 6.82 | 7.76 | 2970 | 43.04 | 43.75 |
| 2022 | 1138 | 75 | 6.59 | 8.44 | 1716 | 22.88 | 8.70 |
| 2023 | 1690 | 120 | 7.10 | 13.50 | 3463 | 28.86 | 60.00 |
| 2024 | 3038 | 200 | 6.58 | 22.50 | 2638 | 13.19 | 66.67 |
| 2025 | 4910 | 305 | 6.21 | 34.32 | 865 | 2.84 | 52.50 |
| Total | 13452 | 889 | - | - | 18672 | - | - |

Citation impact varied across the study period. UK CPP was highest in 2018 (97.35), followed by 2017 (64.59), 2020 (51.50), 2021 (43.04), and 2019 (42.94). More recent years had lower CPP values, including 13.19 in 2024 and 2.84 in 2025. Year-on-year UK publication growth was variable, ranging from 8.70% in 2022 to 75.00% in 2019, with renewed increases in 2023 (60.00%), 2024 (66.67%), and 2025 (52.50%).

The combination of rising publication volume and lower recent CPP is consistent with a citation-lag effect, as newer articles have had less time to accumulate citations. The UK-affiliated output therefore appears to have shifted from an earlier low-volume, high-citation-density phase to a more recent high-volume phase with lower short-term citation density.

### 3. Document types

UK-affiliated publications (n = 889) were distributed across several Scopus-indexed document types (Figure 3). Journal articles accounted for the largest share of output (508 publications; 57.14%), followed by conference papers (261; 29.36%) and reviews (60; 6.75%). Other document types included editorials (18; 2.02%), book chapters, letters, notes, books, and short surveys.

**Figure 3.**
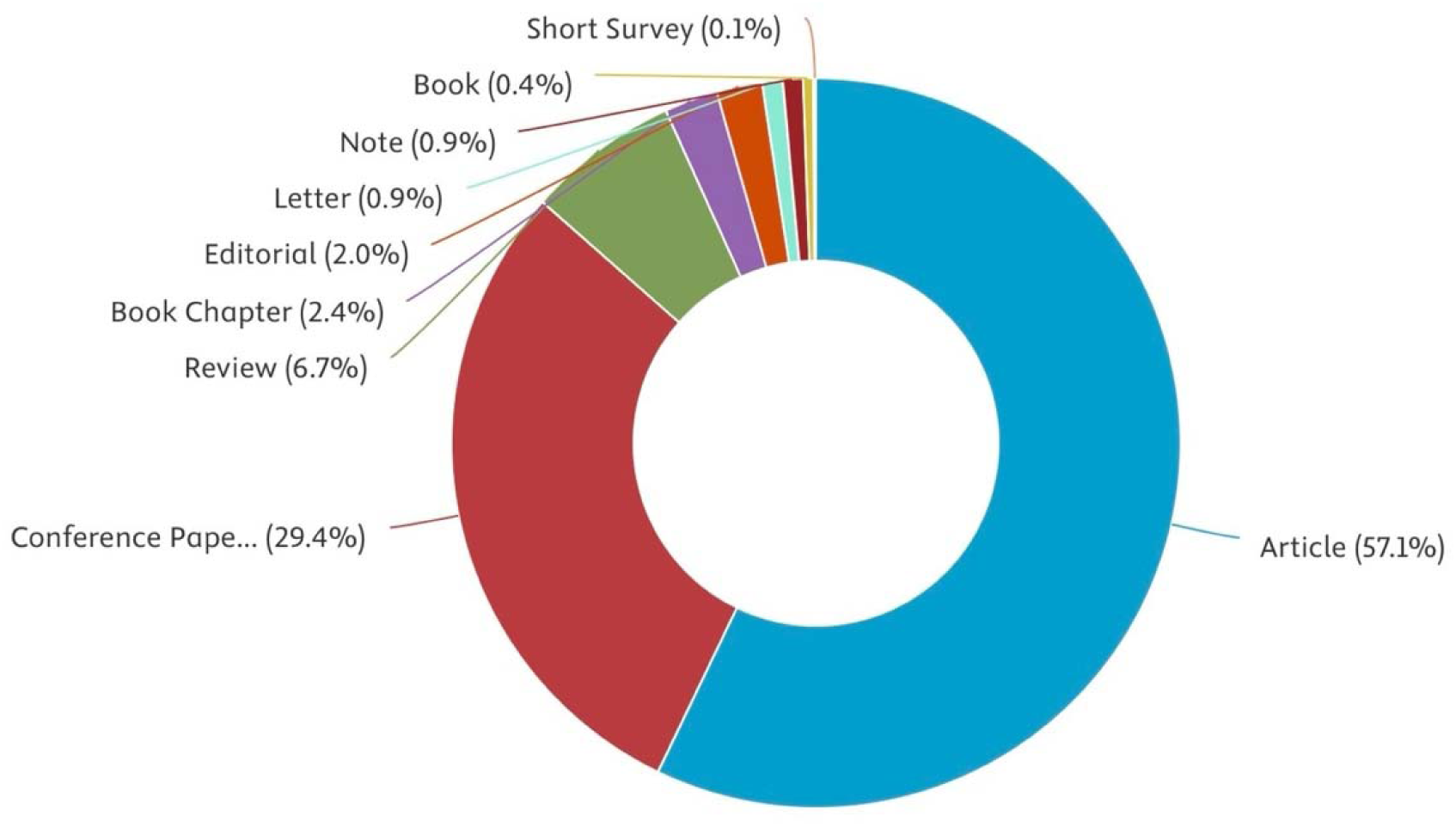
Distribution of document types among United Kingdom-affiliated publications in artificial intelligence applied to medical imaging (n = 889). Proportions are shown for journal articles, conference papers, reviews, and other document types indexed in Scopus.

Comparative document-type distributions across countries showed that the United Kingdom and United States had a higher proportion of journal articles than India and China. The United States had the most journal-dominant profile among the compared countries (65.2% articles; 28.7% conference papers), while the United Kingdom had a majority journal-article profile with a substantial conference-paper component. China had 54.2% journal articles and 31.9% conference papers, while India had the highest conference-paper share among the compared countries (41.9%) and a lower journal-article share (38.4%).

This distribution suggests that the United Kingdom occupies a hybrid dissemination model, positioned between the more journal-dominant United States profile and more conference-heavy models seen in India and, to a lesser extent, China. Countries with higher journal-article proportions, including the United States and United Kingdom, also had higher CPP values than India and China, suggesting that document type may partly contribute to differences in citation visibility.

### 4. Institutional productivity

The nine most productive United Kingdom organisations are presented in Table 3. Imperial College London had the highest publication output (131 publications; 14.74% of UK-affiliated output), followed by King’s College London (110; 12.37%), University College London (101; 11.36%), and the University of Oxford (97; 10.91%). Together, these four institutions accounted for 49.38% of UK-affiliated publications, indicating a substantial concentration of research output within a small number of research-intensive organisations.

**Table 3.** The nine most productive United Kingdom organisations in artificial intelligence in medical imaging research, ranked by total publications. The table reports institutional research output and citation impact measured by total publications (TP), percentage contribution to national output (TP%), total citations (TC), and citations per paper (CPP).

| S. No | Name of organisation | TP | TP (%) | TC | CPP |
| --- | --- | --- | --- | --- | --- |
| Nine most productive organisations |  |  |  |  |  |
| 1 | Imperial College London | 131 | 14.74 | 3310 | 25.27 |
| 2 | King's College London | 110 | 12.37 | 3462 | 31.47 |
| 3 | University College London | 101 | 11.36 | 4171 | 41.30 |
| 4 | University of Oxford | 97 | 10.91 | 3666 | 37.79 |
| 5 | University of Cambridge | 75 | 8.44 | 1689 | 22.52 |
| 6 | University of Oxford Medical Sciences Division | 45 | 5.06 | 1642 | 36.49 |
| 7 | The University of Edinburgh | 38 | 4.27 | 1254 | 33.00 |
| 8 | National Heart and Lung Institute | 37 | 4.16 | 1713 | 46.30 |
| 9 | University of Leeds | 33 | 3.71 | 721 | 21.85 |
| TP: Total papers, TC: Total citations, CPP: Citations per paper |  |  |  |  |  |

Citation impact varied across institutions. The National Heart and Lung Institute had the highest CPP among the listed organisations (46.30), followed by University College London (41.30), the University of Oxford (37.79), the University of Oxford Medical Sciences Division (36.49), and the University of Edinburgh (33.00). Imperial College London had the highest publication volume but a lower CPP (25.27) than several lower-output institutions.

These findings indicate that institutional productivity and citation efficiency did not always move together. Some institutions produced large volumes of research, while others achieved higher average citation impact despite lower publication counts. This suggests that institutional influence in the dataset was shaped by both publication volume and the citation performance of specific outputs.

### 5. Author productivity and collaboration

A total of 3,942 authors contributed to UK-affiliated publications during the study period. The ten most productive authors are shown in Table 4. Yang G. was the most prolific author in the dataset, with 42 publications, 1,965 citations, a CPP of 46.79, and the highest total link strength (TLS 154). Other highly productive authors included Schnabel J.A. (16 publications), Frangi A.F. (14), Rueckert D. (14), Ourselin S. (13), and Tsaftaris S.A. (13).

**Table 4.** The ten most productive authors in United Kingdom-affiliated artificial intelligence in medical imaging research and their collaboration strength. The table reports author-level publication output and impact measured by total publications (TP), percentage contribution to total output (TP%), total citations (TC), citations per paper (CPP), and total link strength (TLS) derived from co-authorship network analysis.

| S. No | Name of author | Name of organisation | TP | TP% | TC | CPP | TLS |
| --- | --- | --- | --- | --- | --- | --- | --- |
| 1 | Yang, G. | Imperial College London | 42 | 4.72 | 1965 | 46.79 | 154 |
| 2 | Schnabel, J.A. | King's College London | 16 | 1.80 | 81 | 5.06 | 57 |
| 3 | Frangi, A.F. | University of Manchester | 14 | 1.57 | 96 | 6.86 | 52 |
| 4 | Rueckert, D. | Imperial College London | 14 | 1.57 | 90 | 6.43 | 24 |
| 5 | Ourselin, S. | King's College London | 13 | 1.46 | 847 | 65.15 | 41 |
| 6 | Tsaftaris, S.A. | University of Edinburgh | 13 | 1.46 | 496 | 38.15 | 28 |
| 7 | Alexander, D.C. | University College London | 11 | 1.24 | 1114 | 101.27 | 15 |
| 8 | Botchu, R. | The Royal Orthopaedic Hospital NHS Foundation Trust | 11 | 1.24 | 208 | 18.91 | 25 |
| 9 | Huang, J. | National Heart and Lung Institute | 11 | 1.24 | 280 | 25.45 | 72 |
| 10 | Iyengar, K.P. | Southport and Ormskirk Hospital NHS Trust | 11 | 1.24 | 208 | 18.91 | 25 |

TP: Total papers, TC: Total citation, CPP: Citations per paper, TLS: Total link strength

Citation impact differed substantially among the most productive authors. Alexander D.C. had the highest CPP among this group (101.27), followed by Ourselin S. (65.15), Yang G. (46.79), and Tsaftaris S.A. (38.15). Authors with the highest TLS values are reported in Table 5. Yang G. had the highest TLS (154), followed by Wang C. (79), Huang J. (72), Zhang Y. (62), Schnabel J.A. (57), Pinaya W.H.L. (54), Frangi A.F. (52), Zhang J. (51), Hu Y. (48), and Cardoso M.J. (45).

**Table 5.** Top 10 Most influential authors in the co-authorship network based on collaboration strength in artificial intelligence in medical imaging research. The table ranks authors by total link strength (TLS), reflecting the intensity of co-authorship connections derived from network analysis.

| S. No | Author | TP | TLS |
| --- | --- | --- | --- |
| 1 | Yang, G. | 42 | 154 |
| 2 | Wang, C. | 7 | 79 |
| 3 | Huang, J. | 11 | 72 |
| 4 | Zhang, Y. | 8 | 62 |
| 5 | Schnabel, J.A. | 16 | 57 |
| 6 | Pinaya, W.H.L. | 9 | 54 |
| 7 | Frangi, A.F. | 14 | 52 |
| 8 | Zhang, J. | 13 | 51 |
| 9 | Hu, Y. | 8 | 48 |
| 10 | Cardoso, M.J. | 8 | 45 |
| TP: Total papers, TLS: Total link strength |  |  |  |

For co-authorship analysis, 327 authors met the minimum threshold of three publications, from which the top 50 authors by TLS were visualised (Figure 4). The resulting network comprised five clusters, 222 links, and a cumulative TLS of 449. The clusters corresponded broadly to groups working in medical image analysis and neuroimaging; computational imaging and machine learning; GAN-based reconstruction and image-to-image translation; stroke and neurovascular imaging; and cardiovascular or physiological imaging.

**Figure 4.**
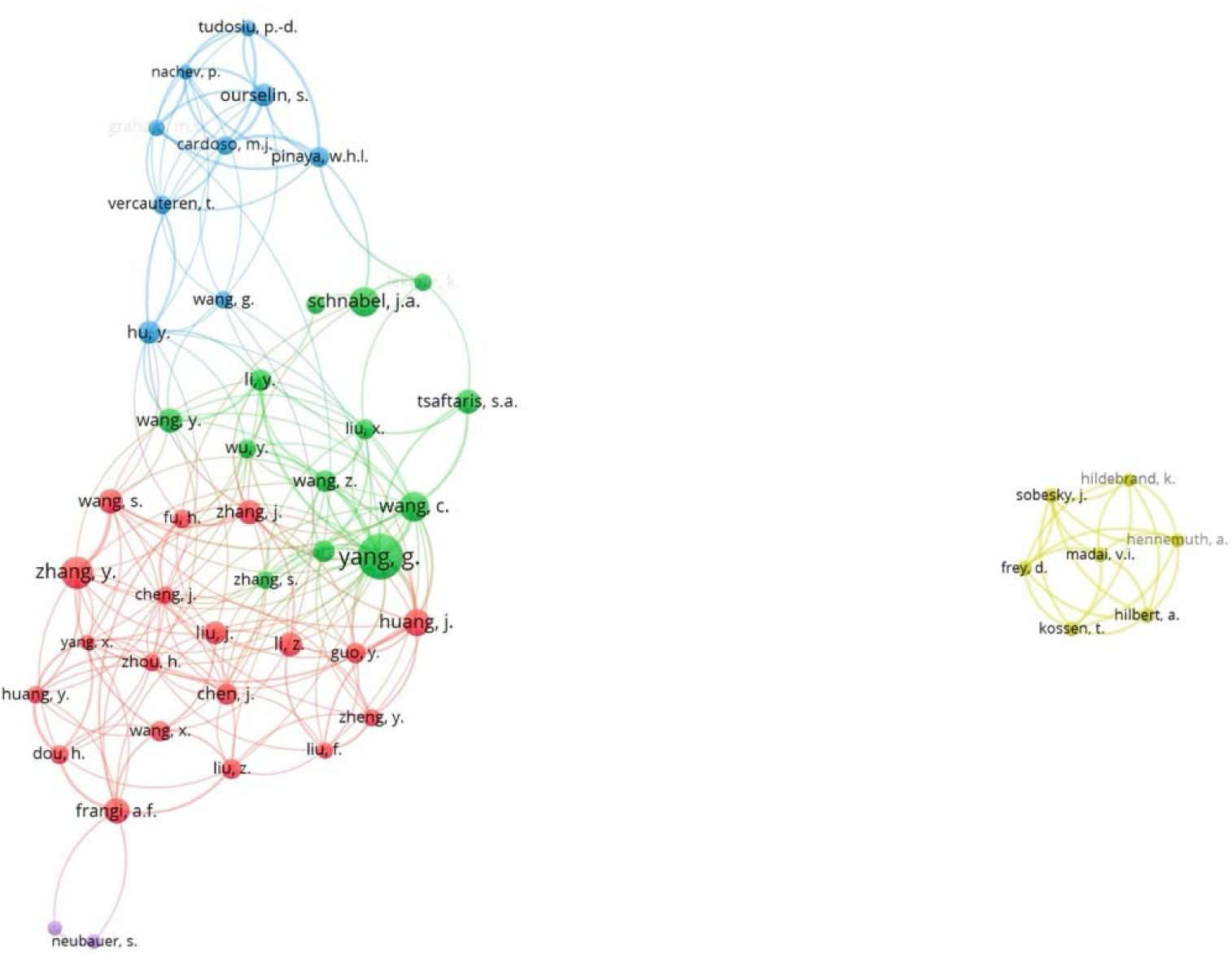
Co-authorship network of authors publishing on artificial intelligence in medical imaging, with emphasis on generative and foundation-based methods, visualised using VOSviewer (v1.6.19). Node size represents publication volume, link thickness indicates co-authorship strength, and colours denote collaboration clusters identified through network analysis.

The author-level findings show a networked research landscape with several interconnected hubs rather than a single dominant group. High TLS values among selected authors suggest that collaboration is an important feature of UK-affiliated AI imaging research, particularly in areas requiring technical, clinical, and methodological expertise.

### 6. External funding

Of the 889 UK-affiliated publications, 552 (62.09%) reported at least one external funding source. A total of 160 distinct funding organisations were identified. The three most frequently reported funding bodies accounted for 33.30% of funded UK-affiliated publications, indicating concentration of funding among a small number of major agencies.

The leading funding organisations are shown in Figure 5. The Engineering and Physical Sciences Research Council (EPSRC) was the most frequently acknowledged funder, with approximately 125 funded documents. Other frequently acknowledged funders included the National Natural Science Foundation of China (approximately 85-90 documents), UK Research and Innovation (more than 80), Horizon 2020 Framework Programme (more than 70), Wellcome Trust (approximately 70), and the Medical Research Council (approximately 60). International funders, including the National Institutes of Health and the European Research Council, were also represented.

**Figure 5.**
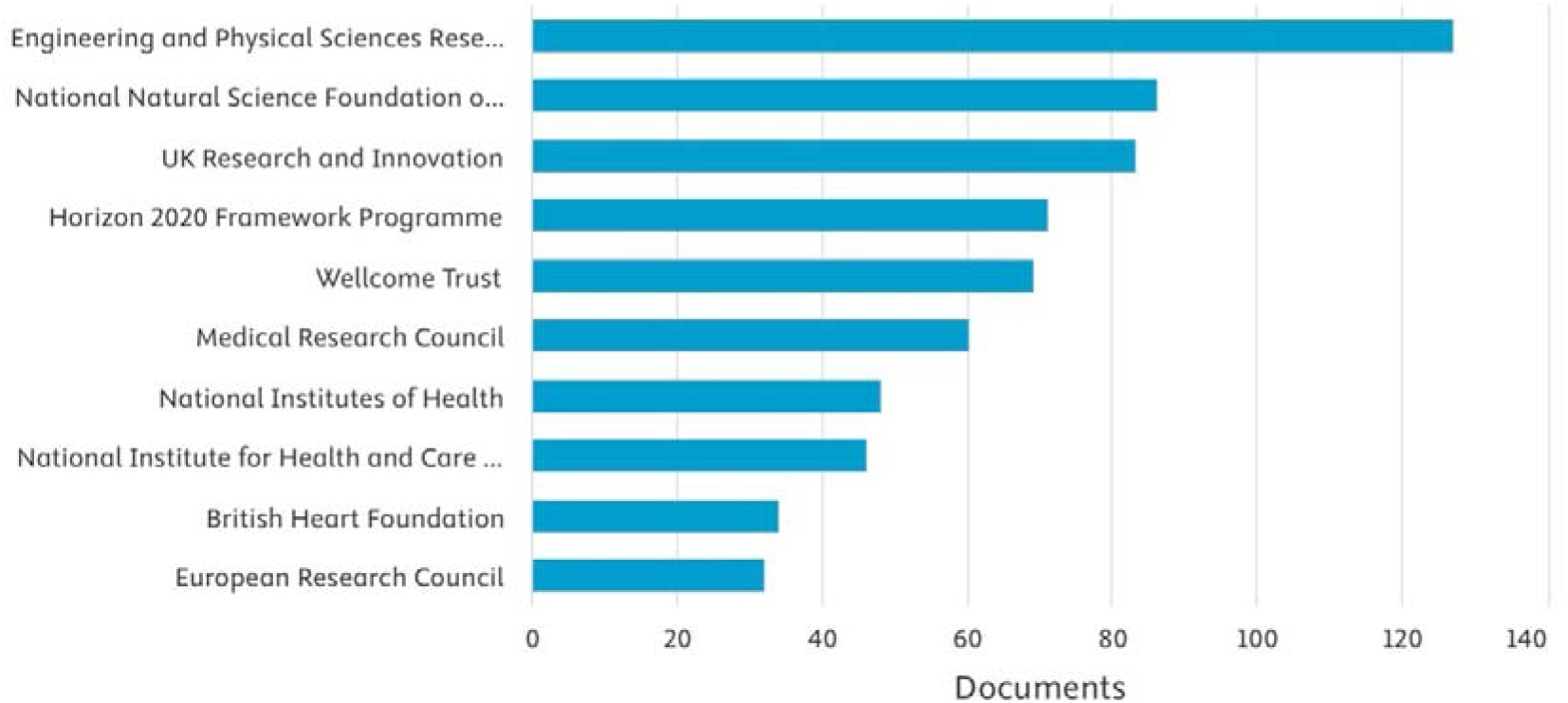
Leading funding organisations acknowledged in United Kingdom-affiliated publications on artificial intelligence in medical imaging. Bars represent the number of Scopus-indexed documents reporting financial support from each funding body.

This funding profile indicates that UK-affiliated AI imaging research in this dataset was supported by a mixture of engineering, computational science, biomedical, and international funding sources. The prominence of EPSRC and UKRI suggests a strong technical and computational base, while the presence of Wellcome Trust, MRC, NIH, and ERC funding reflects biomedical and international dimensions of the research landscape.

### 7. Leading publishing journals

UK-affiliated publications were distributed across 160 journals. The top 20 journals published 337 papers, accounting for 37.91% of all UK journal articles in the dataset. Lecture Notes in Computer Science recorded the highest publication count, with 93 papers. Citation impact varied across journals, with NeuroImage (CPP 76.7) and IEEE Transactions on Medical Imaging (CPP 73.45) among the journals with the highest CPP values.

The journal distribution reflects the interdisciplinary nature of AI in medical imaging, with outputs appearing across computer science, engineering, imaging, neuroscience, and clinical journals. The presence of technically oriented venues among the most productive and highly cited outlets is consistent with the methodological emphasis of generative and foundation-based AI imaging research.

### 8. Keyword analysis

A total of 7,685 keywords were identified across UK-affiliated publications. After applying a minimum occurrence threshold of 10, 332 keywords met the threshold. Following exclusion of generic indexing terms, 40 keywords were retained for network analysis. The most frequently occurring keywords were “deep learning” (291 occurrences), “medical imaging” (264), “diagnostic imaging” (243), and “generative adversarial networks” (204).

Keyword co-occurrence analysis identified three principal clusters (Figure 6). The first cluster was centred on generative and deep learning methodologies and included terms such as generative adversarial networks, adversarial networks, convolutional neural networks, deep learning, image reconstruction, image enhancement, and image segmentation. The second cluster was MRI- and diffusion-imaging-focused and included terms such as magnetic resonance imaging, diffusion, diffusion weighted imaging, diffusion magnetic resonance imaging, neuroimaging, and signal-to-noise ratio. The third cluster related to broader artificial intelligence and diagnostic imaging workflows and included artificial intelligence, machine learning, diagnostic imaging, computed tomography, image analysis, radiology, and large language model.

**Figure 6.**
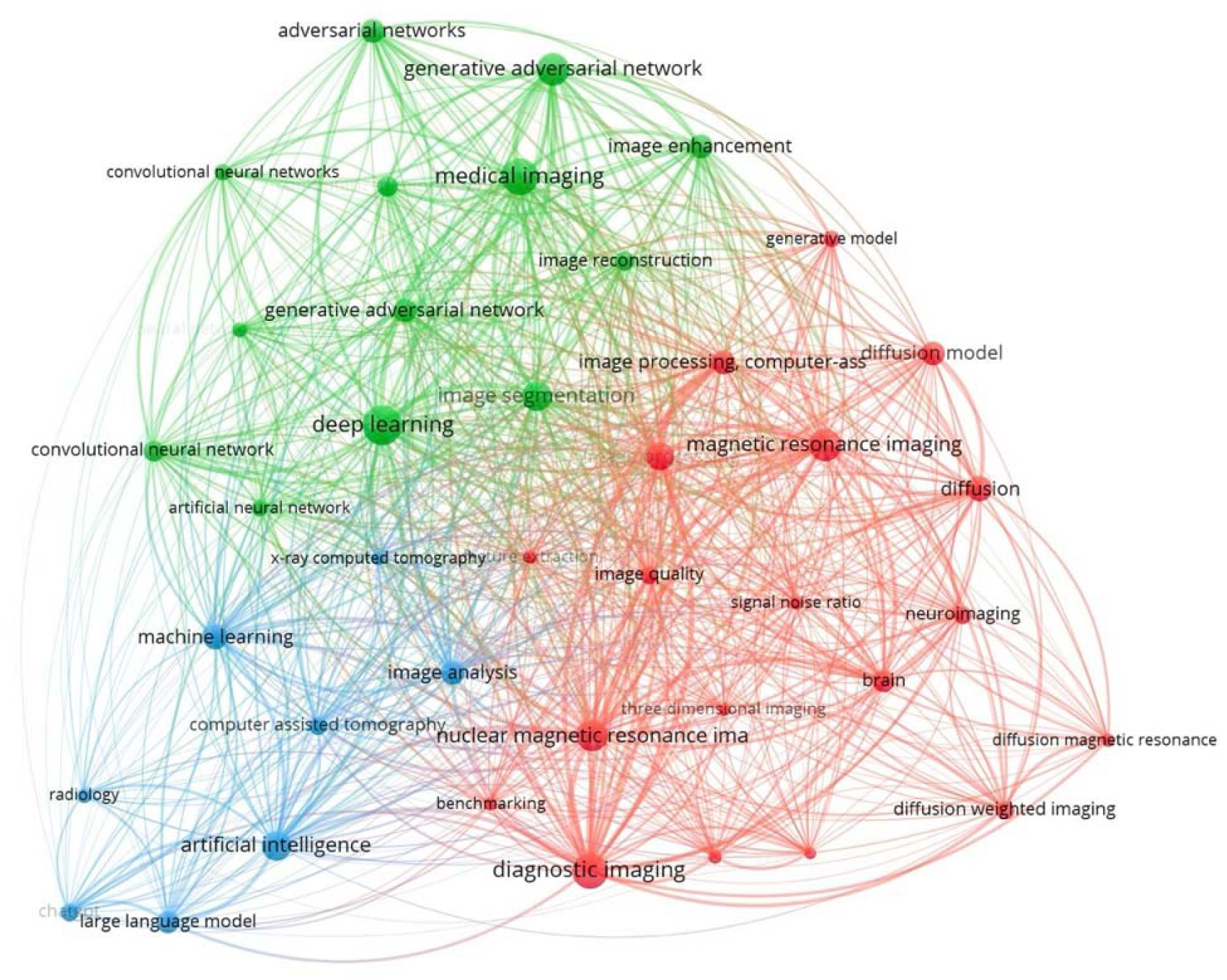
Keyword co-occurrence network of United Kingdom-affiliated publications in artificial intelligence applied to medical imaging, visualised using VOSviewer (v1.6.19). Node size reflects keyword frequency, link thickness represents co-occurrence strength, and colours indicate thematic clusters identified through network analysis.

These clusters suggest that UK-affiliated work in this dataset is organised around both technical model development and imaging-specific applications. The prominence of GANs, reconstruction, enhancement, segmentation, MRI, diffusion, and large language models indicates a research landscape spanning earlier generative modelling approaches and newer foundation-model or multimodal directions.

### 9. Highly cited publications

The 20 most highly cited UK-affiliated publications are presented in Table 6. Together, these papers accrued 5,983 citations, with individual citation counts ranging from 116 to 923. Average citations per year ranged from 16.57 to 164. Publication years ranged from 2017 to 2024, with most highly cited papers published from 2019 onwards.

**Table 6.** The 20 most highly cited United Kingdom-affiliated publications in artificial intelligence applied to medical imaging, with emphasis on generative and foundation-based methods. The table reports total citations, average citations per year, and average citations per year during the first five years following publication, alongside publication year and first author.

| Rank | Citations | Average citations per year | Average citations per year in the first 5 years since publication | First Author | Year | Title |
| --- | --- | --- | --- | --- | --- | --- |
| 1 | 923 | 115.38 | 138.6 | Yang, G. | 2018 | DAGAN: Deep De-Aliasing Generative Adversarial Networks for Fast Compressed Sensing MRI Reconstruction<sup>[14]</sup> |
| 2 | 492 | 164 | N/A | Zhou, Y. | 2023 | A foundation model for generalizable disease detection from retinal images<sup>[18]</sup> |
| 3 | 508 | 84.67 | 97 | Armanious, K. | 2020 | MedGAN: Medical image translation using GANs<sup>[15]</sup> |
| 4 | 478 | 59.75 | 58 | Gibson, E. | 2018 | NiftyNet: a deep-learning platform for medical imaging<sup>[16]</sup> |
| 5 | 364 | 60.67 | 71.4 | Kazeminia, S. | 2020 | GANs for medical image analysis<sup>[17]</sup> |
| 6 | 297 | 148.5 | N/A | Huang, Y. | 2024 | Segment anything model for medical images?<sup>[19]</sup> |
| 7 | 271 | 67.75 | N/A | Wyatt, J | 2022 | AnoDDPM: Anomaly Detection with Denoising Diffusion Probabilistic Models using Simplex Noise<sup>[20]</sup> |
| 8 | 218 | 54.5 | N/A | Pinaya, W.H.L | 2022 | Brain Imaging Generation with Latent Diffusion Models<sup>[21]</sup> |
| 9 | 193 | 64.33 | N/A | Jiang, X. | 2023 | Deep Learning Generation for Medical Image-Based Cancer Diagnosis<sup>[37]</sup> |
| 10 | 202 | 28.86 | 36.2 | Han, C. | 2019 | Combining noise-to-image and image-to-image GAN's: Brain MR Image augmentation for tumour detection<sup>[38]</sup> |
| 11 | 183 | 26.14 | 30.4 | Ben-Cohen, A. | 2019 | Cross-modality synthesis from CT to PET using FCN and GAN networks for improved automated lesion detection<sup>[39]</sup> |
| 12 | 170 | 34 | 34 | Han, C. | 2021 | MADGAN: unsupervised medical anomaly detection GAN using multiple adjacent brain MRI slice reconstruction<sup>[40]</sup> |
| 13 | 163 | 32.6 | 32.6 | Ma, Y. | 2021 | Structure and Illumination Constrained GAN for Medical Image Enhancement<sup>[41]</sup> |
| 14 | 165 | 18.33 | 22 | Chartsias, A. | 2017 | Adversarial image synthesis for unpaired multi-modal cardiac data<sup>[42]</sup> |
| 15 | 138 | 69 | N/A | Wu, J. | 2024 | MedSegDiff-V2: Diffusion-Based Medical Image Segmentation with Transformer<sup>[43]</sup> |
| 16 | 144 | 24 | 28.8 | Dar, S.U.H | 2020 | Prior-guided image reconstruction for accelerated multi-contrast MRI via generative adversarial networks<sup>[44]</sup> |
| 17 | 118 | 59 | N/A | Dayarathna, S. | 2024 | Deep learning-based synthesis of MRI, CT and PET: Review and analysis<sup>[45]</sup> |
| 18 | 125 | 17.86 | 22.4 | Rau, A. | 2019 | Implicit domain adaptation with conditional generative adversarial networks for depth prediction in endoscopy<sup>[46]</sup> |
| 19 | 121 | 24.2 | 24.2 | Karakanis,S. | 2021 | Lightweight deep learning models for detecting COVID-19 from chest X-ray images<sup>[47]</sup> |
| 20 | 116 | 16.57 | 19 | Han, C. | 2019 | Synthesizing Diverse Lung Nodules Wherever Massively: 3D Multi-Conditional GAN-Based CT Image Augmentation for Object Detection<sup>[48]</sup> |

The most cited publication was DAGAN: Deep De-Aliasing Generative Adversarial Networks for Fast Compressed Sensing MRI Reconstruction (Yang, 2018), with 923 citations and an average citation rate of 115.38 citations per year.^[14]^ Other highly cited GAN-related publications included MedGAN (508 citations),^[15]^ NiftyNet (478 citations),^[16]^ and GANs for medical image analysis (364 citations).^[17]^ These papers indicate that GAN-based reconstruction, synthesis, augmentation, and image analysis formed an important part of the earlier highly cited literature.

More recent highly cited publications included A foundation model for generalizable disease detection from retinal images (Zhou, 2023), which accrued 492 citations and 164 citations per year,^[18]^ and Segment Anything Model for Medical Images? (Huang, 2024), which accrued 297 citations and 148.5 citations per year.^[19]^ Diffusion-based publications were also represented among the highly cited set, including AnoDDPM (271 citations; 67.75 citations per year)^[20]^ and Brain Imaging Generation with Latent Diffusion Models (218 citations; 54.5 citations per year).^[21]^

Across the top-cited set, MRI-related applications were common, particularly fast reconstruction, image synthesis, augmentation, and anomaly detection. Other represented areas included cross-modality synthesis, cardiac multimodal imaging, retinal imaging, segmentation, and chest X-ray-based COVID-19 detection. The distribution of highly cited papers suggests a temporal movement from GAN-heavy work toward diffusion and foundation-model architectures, although longer citation windows will be required to determine whether this pattern persists.

## DISCUSSION

This bibliometric analysis mapped global and United Kingdom-affiliated research on artificial intelligence in medical imaging between 2017 and 2025, with emphasis on generative and foundation-based methods. The findings show rapid growth in global publication output, a substantial UK-affiliated contribution, relatively high UK citation efficiency, concentrated institutional activity, networked author collaboration, and thematic clustering around generative/deep learning methods, MRI- and diffusion-related applications, and broader diagnostic imaging workflows.

The sustained increase in publication volume mirrors broader bibliometric analyses showing rapid expansion of AI in medicine and radiology since 2019.^[10]^ Within this growth, the present findings indicate that generative and deep learning methods have become prominent themes within medical imaging research. This is reflected by the high frequency of keywords such as “deep learning,” “medical imaging,” “diagnostic imaging,” and “generative adversarial networks,” as well as by the presence of GAN-, diffusion-, and foundation-model-related papers among the most highly cited UK-affiliated publications. These findings are consistent with previous analyses showing that deep learning dominates highly cited AI-in-radiology literature, with neuroimaging and oncology frequently represented among high-citation domains.^[22]^

The United Kingdom ranked fourth globally by publication volume but had a CPP of 21.00, exceeding several countries with higher or comparable publication volumes. This suggests that UK-affiliated work in this dataset achieved relatively high citation visibility. However, citation visibility should not be equated with methodological quality, clinical validity, or real-world implementation. Citation metrics are influenced by publication age, document type, journal visibility, field size, collaboration patterns, and citation practices. The UK’s position is therefore best interpreted as evidence of strong scholarly visibility rather than direct evidence of superior research quality or clinical impact.

The temporal trend in UK-affiliated publications is consistent with citation-lag effects. Earlier years had lower publication volume but substantially higher CPP, while recent years had much higher publication volume and lower short-term citation density. This pattern is expected in fast-moving research areas, where newly published papers have had limited time to accrue citations. The lower CPP in 2024 and 2025 should therefore be interpreted cautiously and should not be taken as evidence of declining influence.

Institutional and author-level analyses showed that UK-affiliated research activity is concentrated within a relatively small number of research-intensive organisations. The top four institutions accounted for nearly half of UK-affiliated output, while citation efficiency varied across organisations. This indicates that publication volume alone does not explain citation impact. Dense co-authorship networks and high total link strength among selected authors further suggest that collaboration is a major feature of this research ecosystem. This is unsurprising in AI medical imaging, where model development, imaging data access, clinical framing, validation, and deployment considerations often require multidisciplinary expertise.

The document-type analysis showed a predominantly journal-based UK dissemination profile, although conference papers also formed a substantial component. This hybrid profile reflects the position of medical imaging AI between clinical radiology, biomedical engineering, and computer science. The association between journal-dominant output and higher CPP should be interpreted cautiously, but it is plausible that journal articles in clinical and imaging venues receive broader citation visibility than conference proceedings. This may partly explain why the United Kingdom and United States, both with higher journal-article proportions than India and China, showed relatively higher CPP values.

Funding patterns suggest that UK-affiliated AI imaging research is supported by a mixture of engineering, computational science, biomedical, and international funding sources. The prominence of EPSRC and UKRI reflects the technical foundation of this field, while the presence of Wellcome Trust, MRC, NIH, Horizon 2020, and ERC funding highlights biomedical and international dimensions. This pattern supports the view that generative and foundation-based AI imaging research is not purely clinical or purely computational, but depends on sustained interdisciplinary funding.

Keyword and highly cited publication analyses suggest thematic evolution over time. Earlier highly cited work was strongly represented by GAN-based image reconstruction, synthesis, augmentation, and anomaly detection, particularly in MRI. More recent highly cited publications included diffusion-based models, foundation models, and general-purpose segmentation approaches. This suggests that the citation landscape is moving from earlier GAN-dominant work toward broader generative and foundation-model architectures. However, because newer publications have shorter citation windows, this apparent shift should be monitored over time rather than treated as definitive.

The prominence of MRI-related applications among highly cited papers may reflect the technical suitability of MRI for reconstruction, denoising, synthesis, acceleration, and image-quality enhancement. MRI also has strong clinical relevance because acquisition time, image quality, and workflow efficiency remain important practical challenges. Nevertheless, bibliometric data cannot determine whether these methods have entered routine clinical workflows, improved diagnostic performance, reduced waiting times, or improved patient outcomes. The observed thematic focus should therefore be interpreted as evidence of research attention and citation visibility, not evidence of clinical adoption.

### Policy implications

Although this study demonstrates rapid bibliometric growth, bibliometric data cannot establish whether AI methods have entered routine NHS practice or delivered real-world clinical benefit. Early studies in accelerated MRI reconstruction, denoising, synthetic data generation, segmentation, and image enhancement describe applications with potential workflow relevance in the context of workforce shortages and rising imaging demand.^[24,25]^ However, implementation-focused studies appear less prominent than methodological publications, suggesting that further prospective evaluation, deployment research, and implementation science are needed before routine clinical translation can be inferred.^[26,27]^

Several barriers remain relevant to implementation, including data governance, interoperability, computational infrastructure, procurement, regulatory evaluation, model monitoring, and accountability.^[28–31]^ Generative imaging models raise additional concerns related to image authenticity, artefact introduction, synthetic data provenance, and clinical responsibility.^[32–34]^ These considerations support the need for standardised evaluation frameworks, transparent reporting, prospective validation, and post-deployment monitoring. Reporting guidance such as CONSORT-AI and TRIPOD+AI may help strengthen future evaluation and reporting of AI-based imaging tools.^[35,36]^

For the UK specifically, the concentration of output in major academic centres and the presence of dense collaboration networks may provide a useful basis for multicentre validation studies, shared infrastructure, and coordinated evaluation frameworks. Priority areas suggested by the bibliometric patterns include MRI reconstruction, image enhancement, segmentation, multimodal imaging, and foundation-model applications. However, future research priorities should be driven not only by citation visibility but also by clinical need, patient benefit, workflow integration, safety, cost-effectiveness, and equity.

### Limitations

This study has several limitations inherent to bibliometric analyses. The analysis was restricted to publications indexed in Scopus and may not capture relevant records indexed exclusively in other databases. Scopus was selected because of its broad coverage of engineering, computer science, biomedical, medical imaging, and conference proceedings literature, which are central dissemination routes for AI research in medical imaging. Although adding databases such as Web of Science or PubMed may increase record capture, cross-database merging introduces challenges including metadata inconsistencies, duplicate records, and discordant citation counts arising from database-specific indexing systems. Accordingly, a single-database approach was adopted to preserve internal consistency and reproducibility of bibliometric indicators.

The findings are also dependent on the chosen search strategy. Although the search was designed to capture generative and foundation-based AI methods in medical imaging, terminology in this field is heterogeneous and rapidly evolving. Some relevant publications may therefore have been missed, while some broader AI imaging studies may have been captured. Citation-based metrics reflect scholarly attention rather than methodological quality, clinical validity, or real-world impact and are influenced by citation-lag effects, particularly for recent publications. Citation counts may also be affected by journal visibility, article type, self-citation, field size, collaboration structure, and publication age.

The study relied on bibliographic metadata and did not assess the technical performance, clinical implementation, regulatory status, ethical characteristics, or patient-level impact of individual AI models. Full counting rather than fractional counting was applied, which may overrepresent contributions from multi-authored or multi-institutional publications. Affiliation-based filtering may also underrepresent UK researchers contributing to international collaborations where UK affiliations were not indexed. Finally, keyword co-occurrence and co-authorship maps are sensitive to threshold choices, author name disambiguation, indexing quality, and VOSviewer settings.

## CONCLUSION

This bibliometric analysis demonstrates rapid global growth in artificial intelligence research applied to medical imaging between 2017 and 2025, with UK-affiliated work representing a substantial and highly cited component of the field. The United Kingdom ranked fourth by publication volume and showed relatively high citation visibility, with research activity concentrated among major academic centres and supported by interdisciplinary funding. Keyword and highly cited publication analyses indicate prominent themes around generative and deep learning methods, MRI reconstruction and enhancement, diffusion-based approaches, and foundation-model architectures. These findings provide a reproducible baseline for monitoring the evolution of AI medical imaging research, while recognising that citation-based indicators do not directly measure clinical implementation, methodological quality, or patient-level impact.

## Ethics approval and consent to participate

Not applicable.

## Consent for Publication

Not applicable

## Availability of Data and Materials

The Scopus bibliographic records retrieved for this study and the cleaned dataset used for analysis can be found here 10.5281/zenodo.18659244

## Author Contributions

J.N. conceived and designed the study, developed the search strategy. J.N. and V.B. performed the literature search and bibliometric analyses, curated and analysed the data, interpreted the results, and drafted and revised the manuscript. The authors read and approved the final manuscript.

## Data Availability

https://zenodo.org/records/18659245

